# Association Between Torque Teno Virus and Systemic Immunodeficiency in Patients With Uveitis With a Suspected Infectious Etiology

**DOI:** 10.1101/2023.01.20.23284502

**Authors:** Ai Fujita Sajiki, Yoshito Koyanagi, Hiroaki Ushida, Kenichi Kawano, Kosuke Fujita, Daishi Okuda, Mitsuki Kawabe, Kazuhisa Yamada, Ayana Suzumura, Shu Kachi, Hiroki Kaneko, Hiroyuki Komatsu, Yoshihiko Usui, Hiroshi Goto, Koji M. Nishiguchi

**Affiliations:** Department of Ophthalmology, Nagoya University Graduate School of Medicine, Nagoya, Japan; Department of Ophthalmology, Yokkaichi Municipal Hospital, Yokkaichi, Japan; Shohzankai Medical Foundation, Miyake Eye Hospital, Nagoya, Japan; Department of Ophthalmology, Tokyo Medical University, Tokyo, Japan

**Author notes:** Co–corresponding author. **Contact information for corresponding author:** Ai Fujita Sajiki, Department of Ophthalmology, Nagoya University Graduate School of Medicine, 65 Tsurumai-cho, Showa-ku, Nagoya 466-8550, Japan. Co–first author (Ai Fujita Sajiki, Yoshito Koyanagi). This study was presented in part at the 126th Annual Meeting of the Japanese Ophthalmological Society in April 2022 and will be presented at the 76th Annual Congress of Japan Clinical Ophthalmology in October 2022.

## Abstract

**Importance:** Torque teno virus positivity in the aqueous humor of uveitis patients could be associated with systemic immunodeficiency.

**Objective:** We explored the correlation between the presence of torque teno virus in the aqueous humor of uveitis patients and clinical information, including immunodeficiency history.

**Design:** This was a retrospective, cross-sectional study.

**Setting:** All participants were recruited at Nagoya University Hospital, Tokyo Medical University Hospital, or Yokkaichi Municipal Hospital between April 2017 and March 2022.

**Participants:** The participants were 58 uveitis patients with a suspected infectious etiology and 24 controls with cataract or age-related macular degeneration.

**Main Outcomes and Measures:** We used quantitative polymerase chain reaction to test all subjects for torque teno virus and multiplex polymerase chain reaction to test uveitis subjects for common ocular pathogens. When possible, both serum and aqueous humor samples were tested. Ocular torque teno virus positivity was compared with age, sex, and a history of systemic immunodeficiency with logistic analysis.

**Results:** Torque teno virus positivity was found in 7 of 31 cases (23%) with herpetic uveitis, 3 of 27 cases (11%) with nonherpetic uveitis, and 0 of 24 controls (0%). Among patients with herpes infection, positivity for torque teno virus was found in 3 of 7 patients (43%) with cytomegalovirus retinitis, 1 of 12 (8%) patients with iridocyclitis, 1 of 7 patients (14%) with acute retinal necrosis, and 2 of 4 patients (50%) with Epstein–Barr virus–related uveitis. Cytomegalovirus retinitis patients showed a significantly higher rate of ocular torque teno virus infection than controls (*P* = .008). Serum analysis revealed torque teno virus positivity in 9 of 10 cases (90%) with uveitis and in all 8 controls (100%). Age- and gender-adjusted logistic analysis revealed a correlation between ocular torque teno virus positivity and systemic immunodeficiency (*P* = .01), but no correlations between ocular torque teno virus and age, gender, or viral pathogenic type.

**Conclusion and Relevance:** This study found that positivity for ocular torque teno virus was correlated with a clinical history of systemic immunodeficiency. This suggests that ocular torque teno virus is a biomarker of systemic immunity.

**Key points:** *Question:* What is the correlation between torque teno virus (TTV) in the aqueous humor of patients with uveitis and the clinical characteristics of these patients?

*Findings:* In this retrospective, cross-sectional study that included 82 patients, ocular TTV was found to be present in 39% of uveitis patients with underlying immunodeficiency and 8% of patients without underlying immunodeficiency; these findings showed statistical significance.

*Meaning:* In patients with uveitis, the presence of ocular TTV may represent systemic immunodeficiency.

## Introduction

Torque teno virus (TTV) is a nonenveloped, single-stranded DNA virus, which was first isolated in 1997 from the serum of a Japanese patient with posttransfusion hepatitis of unknown etiology.^1-3^ In the literature, a high concordance rate of TTV DNA in blood and feces has been reported,^4^ and this may explain the high TTV positivity ratio in the blood of the general population.^3^ However, as TTV is ubiquitous and is found in water,^5^ air,^6^ and various human tissues,^7^ infection from other routes is also possible. Furthermore, TTV has been detected in healthy people, indicating that it is part of the normal mammalian viral flora.^8^ Although the direct involvement of TTV in the development of disease in humans has not been verified,^9^ increased viral loads of TTV have been detected in posttransplant patients treated with immunosuppressive drugs, and TTV has received attention as a biomarker of immune function.^10,11^ Furthermore, a higher titier of TTV is associated with an increased risk of graft-versus-host disease and infections in posttransplant patients.^12^

In the field of ophthalmology, TTV infection has been detected in intraocular samples of patients with culture-positive and culture-negative postoperative endophthalmitis.^13-15^ The presence of ocular TTV is associated with a requirement for additional surgery in cases of postoperative endophthalmitis,^14,15^ suggesting that it has an indirect impact on ocular disease. Moreover, TTV appears to be ubiquitous around the eyes. It has been reported to be present in the conjunctivae (20%-65%)^16,17^ and tears (33%-67%),^18^ making intraocular infection with the virus via surgical wounds possible.^19^ In addition to endophthalmitis, TTV has been detected in the vitreous fluid of patients with seasonal hyperacute panuveitis (SHAPU),^20^ which is a rapidly progressive, rare type of intraocular inflammation of uncertain cause that has been reported solely in Nepal.^21^ However, TTV infection in more common forms of infectious and noninfectious uveitis has not been studied and, unlike other organs, little is known about ocular TTV infection in relation to the systemic immune status of patients.

This study aims to investigate the presence of TTV in the aqueous humor (AH) of patients with uveitis suspected to have an infectious etiology. We also determined whether the results were correlated with the clinical backgrounds of the patients, including their history of immune deficiency.

## Methods

### Study Design

This was a retrospective, observational case series. We followed the tenets of the Declaration of Helsinki, received approval from the Institutional Review Board of Nagoya University Graduate School of Medicine (2020-0598), and registered the study with the University Hospital Medical Information Network (UMIN000044906). Additionally, we obtained written informed consent from all patients for testing of the samples. All participants were recruited at Nagoya University Hospital, Tokyo Medical University Hospital, or Yokkaichi Municipal Hospital, and patient clinical information was obtained retrospectively from medical records. The STROBE checklist was used for reporting the data collection methods.^22^

### Sample Collection

This study included consecutive patients diagnosed with unilateral or bilateral anterior uveitis or panuveitis based on a comprehensive ophthalmological examination and clinical history. Only eyes that underwent multiplex polymerase chain reaction (mPCR)^23-25^ testing to assess herpes-related infection were analyzed. We used the remaining surplus of the AH samples for mPCR testing in the uveitis patients. Patients were excluded if they had other types of intraocular inflammation, such as postprocedural endophthalmitis or noninfectious uveitis (eg, sarcoidosis, Vogt-Koyanagi-Harada disease, and Behçet disease). Control subjects had cataract or exudative age-related macular degeneration (AMD). AH samples were collected from the cataract patients at the beginning of the procedure, and from the AMD patients at the time of treatment by vitreous injection of anti-vascular endothelial growth factor agents. All samples were collected between April 2017 and March 2022, and were stored at −80°C.

### DNA Purification and Quantitative Polymerase Chain Reaction for TTV

Viral genomic DNA was purified and extracted from either 10-µL samples of the AH or 200-µL samples of serum using the QIAamp MinElute Virus Spin Kit (Qiagen, Hilden, Germany) per the manufacturer’s protocol. Subsequently, quantitative polymerase chain reaction (qPCR) testing was performed for TTV using the TaqMan Fast Advanced Master Mix (Thermo Fisher Scientific, Waltham, MA) on the QuantStudio 5 Real-Time PCR System (Thermo Fisher Scientific). For each reaction, we mixed the following samples and reagents to a final volume of 20 µL: 2 µL of purified DNA, 0.8 µL of primer/probe mix (forward 10 µM, reverse 10 µM, Taqman probe 5 µM), 10 µL of TaqMan Fast Advanced Master Mix (2×), and 7.2 µL of nuclease-free water. Cycling conditions were as follows: a 2-min incubation at 50 °C and 20-s polymerase activation at 95 °C, followed by 1-s denaturation at 95 ° C and 20 s of annealing/extending at 60 °C (repeated for 40 cycles). The primer/probe mix contained the following, all reported previously^26^: forward primer 5’
s-GTGCCGIAGGTGAGTTTA-3’, reverse primer 5’-AGCCCGGCCAGTCC-3’, and probe FAM-5’-TCAAGGGGCAATTCGGGCT-3’-TAMRA. For the standard curve, standard TTV DNA was serially diluted 10 times from 1×10^3^ to 10^12^ copy/mL.

### Statistical Analysis

All statistical analyses were performed using SPSS (v 28; IBM Corp, Armonk, NY), except for the post hoc calculation analysis, which was performed using G*Power (v 3.1.9.6; Heinrich Heine University Düsseldorf, Germany).

Best-corrected visual acuity (BCVA) was recorded as decimal values and converted to logarithm of the minimum angle of resolution (logMAR) values for the statistical analysis. For BCVA of counting fingers or less, logMAR values were assigned as previously described.^27^ Data were presented as numbers and percentages for categorical data and as mean and SD for distributed variables, unless otherwise specified. The normality of the distribution of continuous variables was assessed using the Shapiro–Wilk test. Age, BCVA, and TTV copy numbers were found to be nonnormally distributed. The Fisher exact test was used for categorical variables, and a logistic regression analysis was used for age- and gender-adjusted analysis. A post hoc calculation using α = .05 for the logistic regression analysis showed that the value of 1 – β was .879. Therefore, the detection power was found to be more than 80%.

## Results

### Clinical Characteristics of the Patients

Table 1 presents the clinical characteristics of the enrolled subjects. Among 58 uveitis patients suspected of having an infectious etiology, mPCR testing was positive in 31 cases and negative in 27 cases. This study included 24 nonuveitis control subjects, including 15 with cataract (63%) and 9 with AMD (38%). Types of uveitis detected in the mPCR-positive and mPCR-negative patients included, respectively, panuveitis (16/31, 52%; 10/27, 37%), anterior uveitis (14/31, 45%; 14/27, 52%), and posterior uveitis (1/31, 3%; 1/27, 4%). Among 31 patients with herpetic pathogens, 2 cases of herpes simplex virus (HSV)-1 (7%), 2 cases of HSV-2 (7%), 14 cases of varicella–zoster virus (VZV; 45%), 4 cases of EBV (13%), and 9 cases of cytomegalovirus (CMV; 29%) were identified. Positive histories of immunodeficiency were observed in 42% (13/31), 19% (5/27), and 21% (5/24) of patients in the mPCR-positive, mPCR-negative, and control groups, respectively. The inconsistency with the calculations for immunodeficiency below arose because 3 patients each had 2 immunodeficiency conditions.

**Table 1.**
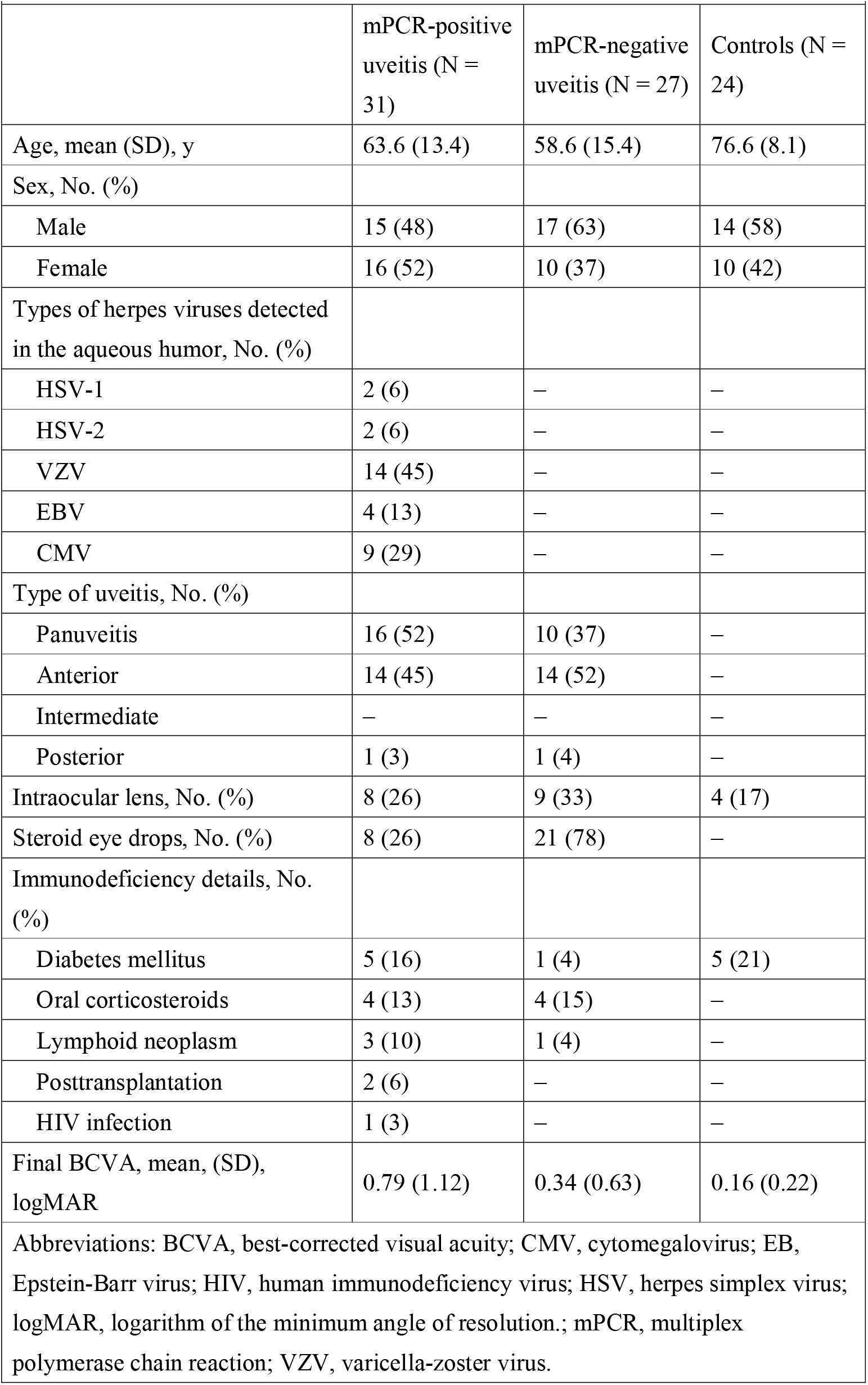
Clinical Characteristics of Controls and Patients With mPCR-Positive Or mPCR-Negative Uveitis

### Detection of TTV in the AH Samples

The qPCR analysis of the AH samples revealed TTV in 23% (7/31) of mPCR-positive uveitis patients, 11% (3/27) of mPCR-negative uveitis patients, and 0% (0/24) of controls. Ocular TTV infection was more frequent in mPCR-positive cases than controls (*P* = .01), but no significant difference was shown between mPCR-negative cases and controls (Figure 1A; eTable 1 in the Supplement). Figure 1B provides details of the diagnoses in cases in which the viral pathogen was detected using mPCR. Comparison of TTV infection rates in controls and patients with a specific diagnosis revealed no significant differences, except for patients with CMV retinitis, who had a higher rate of TTV infection (*P* = .008; eTable 2 in the Supplement).

**Figure 1.**
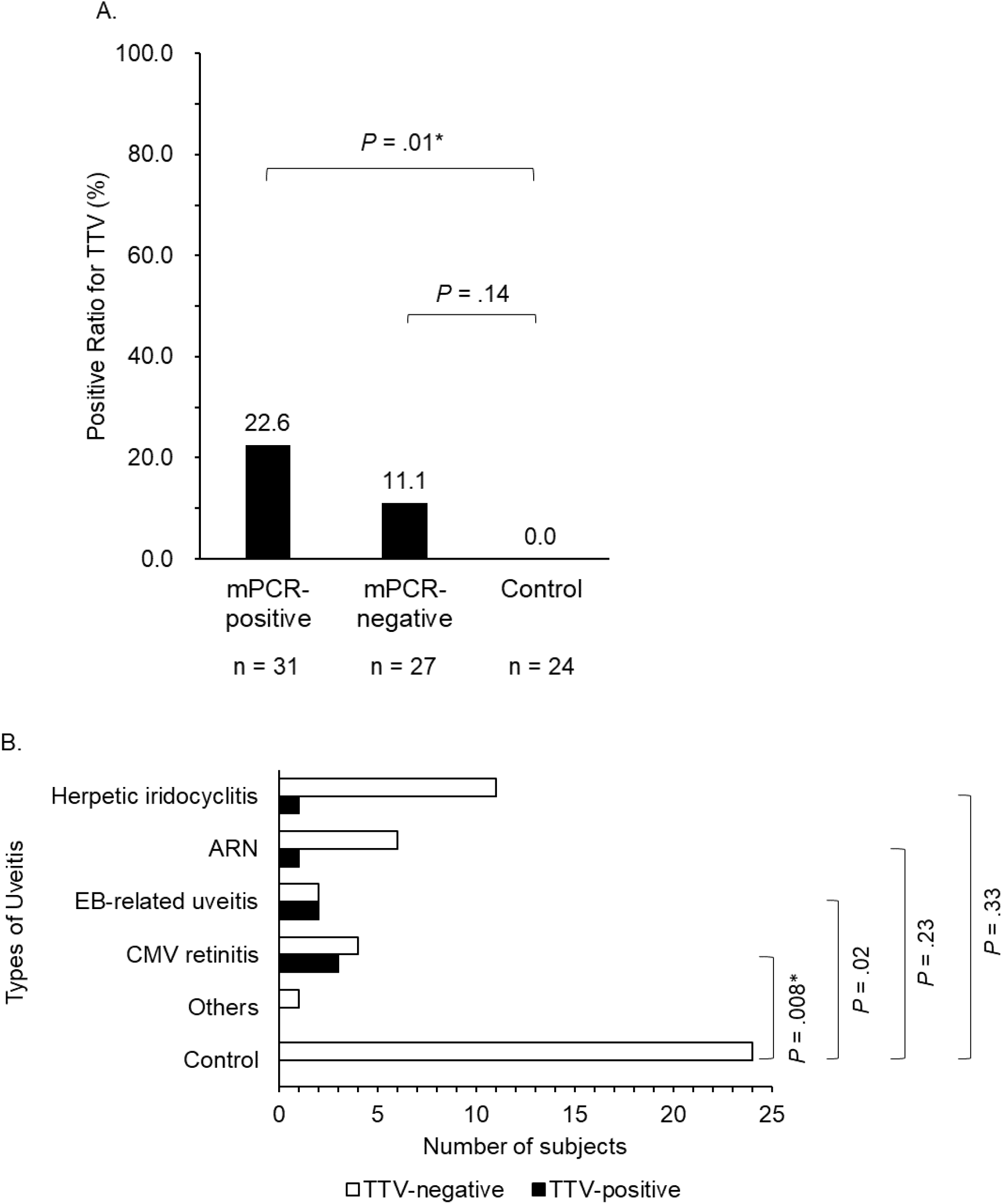
The disease-specific positive ratio for torque teno virus (TTV) in the AH.**A**. The positive ratio for TTV, determined with multiplex PCR (mPCR). Each bar indicates the percentage of patients with a positive ratio for TTV in each group. **B**. Breakdown of mPCR-positive herpetic uveitis patients and controls. Each bar indicates the number of TTV-positive and TTV-negative samples per disease. HSV-1, HSV-2, VZV, and CMV were identified as the underlying pathogens of herpetic iridocyclitis. HSV-1, HSV-2, and VZV were identified as the pathogen of acute retinal necrosis (ARN). The “others” item includes keratouveitis. The Fisher exact probability test was used to calculate *P* values, with *P* < .03 for Fig. 1A and *P* < .01 for Fig. 1B being defined as significant, after Bonferroni correction. Abbreviations: ARN, acute retinal necrosis; CMV, cytomegalovirus; EB, Epstein-Barr virus; mPCR, multiplex polymerase chain reaction; TTV, torque teno virus.

### Assessment of Ocular TTV and Systemic Immunodeficiency

To assess the correlation between systemic immune status and ocular TTV infection in the patients with uveitis, we recategorized the patients into 2 groups based on the presence or absence of a clinical history of underlying systemic immune deficiency. Ocular TTV was found to be more frequent in uveitis patients with underlying immunodeficiency (7/18, 39%) versus those without (3/40, 8%), a statistically significant difference (*P* = .007; Fig. 2, eTable 3 in the Supplement). Furthermore, after applying logistic regression analysis to assess the effect of age and gender on ocular TTV infection, the correlation remained significant (Table 2). Although a similar logistic regression analysis was conducted for the presence of TTV in the AH samples and the type of virus detected by mPCR, no significant correlations were found.

**Table 2.** Age- and Gender-Adjusted Logistic Regression Analysis of Torque Teno Virus in the Aqueous Humor and Immunodeficiency

|  |  |  |
| --- | --- | --- |
|  | Odds ratio (95% CI) | *P* Value |
| Age | 1.026 (0.959 to 1.097) | .45 |
| Sex | 0.778 (0.159 to 3.810) | .76 |
| Immunodeficiency | 8.131 (1.653 to 40.005) | .01* |
| Model $\chi^2$ test: *P* = .03; Hosmer–Lemeshow test: *P* = .59 | | |

**Figure 2.**
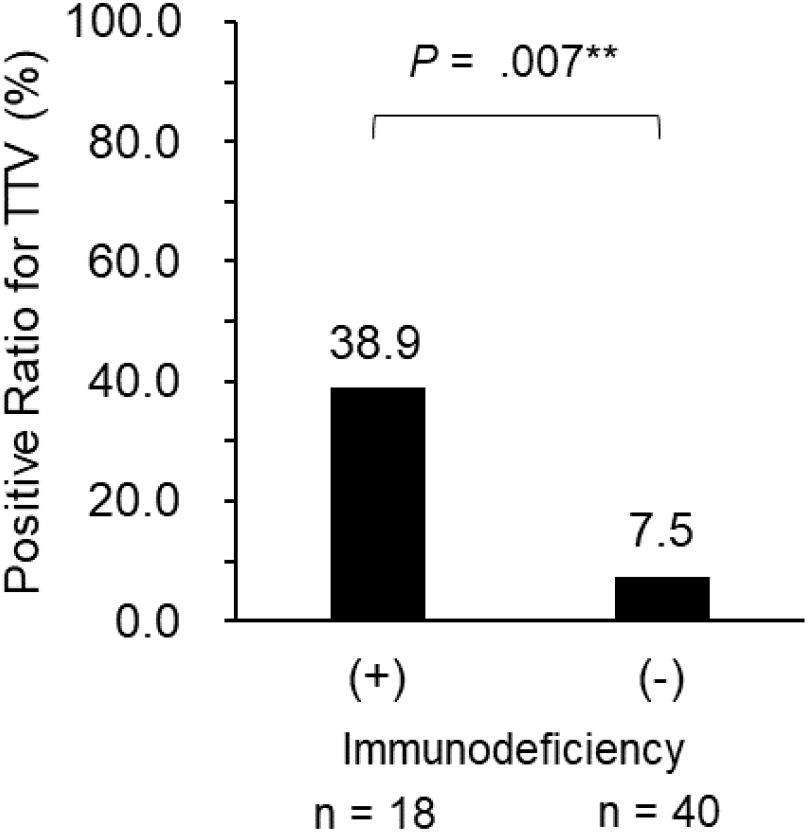
The positive ratio for torque teno virus (TTV) by immunodeficiency status. Each bar indicates the positive ratio for TTV in AH samples from patients with (+) and without (−) immunodeficiency, respectively. The Fisher exact probability test was used to calculate *P* values, with *P* < .05 defined as significant. Abbreviation: TTV, torque teno virus.

For selected cases in which serum samples were obtained prior to the extraction of AH samples, TTV infection was also assessed with qPCR in the serum samples. TTV infection was confirmed in the serum of all mPCR-positive cases (4/4, 100%), nearly all mPCR-negative cases (5/6, 83%), and all controls (8/8, 100%); these differences were not statistically significant (*P* = .56; eFigure in the Supplement). In addition, the TTV copy number in the AH samples was determined in cases with a sufficient remaining quantity of sample. No significant differences in copy number in the serum were detected in patients with and without a history of immunodeficiency (*P* = .35; Mann–Whitney U test, Table 3).

**Table 3.** Copy Numbers of TTV Detected in Samples from Patients With Uveitis

|  |  |  |  |  |
| --- | --- | --- | --- | --- |
| Types of uveitis | Age | Sex | Copy number of TTV in AH (copies/mL) | History of systemic immunodeficiency |
| mPCR-negative uveitis | 60's | Female | 1.54×10<sup>9</sup> | None |
| CMV retinitis | 70's | Male | 4.98×10<sup>6</sup> | Lymphoid neoplasm |
| Herpetic iridocyclitis (VZV) | 60's | Male | 2.38×10<sup>6</sup> | None |
| CMV retinitis | 60's | Male | 1.26×10<sup>6</sup> | Lymphoid neoplasm |
| CMV retinitis | 60's | Female | 1.23×10<sup>6</sup> | Lymphoid neoplasm |
| mPCR-negative uveitis | 70's | Male | 2.11×10<sup>5</sup> | None |
| mPCR-negative uveitis | 40's | Male | 2.25×10<sup>4</sup> | None |
| ARN (HSV-2) | 60's | Female | 6.25×10<sup>3</sup> | Oral corticosteroids |
| EB-related uveitis | 60's | Female | 3.74×10<sup>3</sup> | Post-transplantation and oral corticosteroids |
| EB-related uveitis | 60's | Female | 1.9×10<sup>1</sup> | Diabetes mellitus |
| Abbreviations: AH, aqueous humor; ARN, acute retinal necrosis; CMV, cytomegalovirus; EB, Epstein-Barr virus; HSV, herpes simplex virus; mPCR, multiplex polymerase chain reaction; TTV, torque teno virus; VZV, varicella-zoster virus. |  |  |  |  |

## Discussion

This study detected TTV DNA in the AH of herpetic and nonherpetic uveitis patients but not in the AH of control subjects with cataract or AMD. When we compared the frequency of ocular TTV infection among patients with a specific infectious diagnosis, we found that patients with CMV retinitis had a higher rate of ocular TTV infection. CMV retinitis is usually seen in immunocompromised patients, which agrees well with our finding that there was a higher frequency of ocular TTV infection in patients with underlying systemic immunodeficiencies, such as diabetes mellitus, oral corticosteroid intake, lymphoid neoplasm, post–organ transplantation, and human immunodeficiency virus (HIV) infection. This is in line with of reports on patients with nonocular diseases, who show a correlation between reduced immune status and TTV infection. For example, TTV infection has been found to be more frequent in the serum of children receiving immunosuppressive treatment after liver transplantation than in healthy controls,^28^ and TTV plasma levels have been shown to be higher in HIV-infected patients with poor immune recovery, as indicated by low CD4 levels, even after antiretroviral therapy.^29^ However, the association of ocular TTV infection and systemic immunodeficiency has not been well studied to date. The markedly higher rate of ocular TTV infection in uveitis patients with a clinical history of immunosuppression compared to those without such a history or controls suggests that ocular TTV is a biomarker of systemic immune status. This is in sharp contrast to serum TTV; nearly all patients and controls were found to be infected with TTV. A key related question is whether ocular TTV infection reflects intraocular immune depression, as this has important clinical implications.

We did not detect TTV in our controls with cataract or AMD. However, there are contradictory past reports. Smits et al^20^ showed that TTV was detected in vitreous samples from 91% of patients with SHAPU, 47% with endophthalmitis, 17% with nonbacterial or fungal infectious uveitis, and 0% of controls with retinal detachment. Lee et al^13^ reported that TTV was detected in the vitreous samples of 57% of patients with culture-positive endophthalmitis and 100% of patients with culture-negative endophthalmitis, but in 0% of control subjects with noninflammatory ocular conditions, such as epiretinal membrane and macular hole. In contrast, Naik et al^14^ reported that the vitreous fluid was positive for TTV in 38% of controls, including patients with noninfectious retinal diseases, while Emre et al^18^ detected TTV in 12.5% of AH samples from cataract patients. The seemingly contradictory results reported in the present report and by Naik et al^14^ may have arisen from Naik’s inclusion of control subjects with retinal pathologies, such as proliferative diabetic retinopathy (24.5%), which is associated with immune dysfunction and intraocular inflammation. The reasons for the differences between our results and those of Emre et al^18^ remain unknown. Potential explanations include differences in the sensitivity and specificity of the detection methods, including the primer design,^30^ or in the social and ethnic backgrounds of the subjects.

Although this study provides new insights into the relationship between systemic immunodeficiency and TTV in the AH of patients with uveitis, it has several limitations worth acknowledging. First, we found no direct evidence associating ocular TTV infection and systemic immunodeficiency. To prove this association, a comparison of systemic immune biomarkers and ocular TTV infection will be necessary. Second, the exact role of TTV infection in ocular inflammation remains unclear. A correlation analysis of ocular inflammatory markers and TTV infectious titer in a larger number of samples could potentially address this issue. Alternatively, comparing the presence and amount of ocular TTV with the clinical course of uveitis may provide important clues, as similar approaches have been carried out successfully in ocular and nonocular conditions.^12,15,31^ In addition, as samples were collected from 3 unrelated institutions, the results of clinical examinations could not be recorded under uniform conditions, hampering an accurate phenotypic comparison of TTV-positive and TTV-negative cases. Hence, a larger, prospective validation study is warranted to substantiate our results.

In conclusion, this study demonstrated a significantly higher rate of ocular TTV infection in patients with uveitis with a suspected infectious etiology and in those with evidence of systemic immunodeficiency. Future studies should assess the role of ocular TTV infection in intraocular inflammation and immunity.

## Data Availability

All data produced in the present work are contained in the manuscript.

## Acknowledgments

Ai Fujita Sajiki and Yoshito Koyanagi contributed equally as co–first authors. Ai Fujita Sajiki and Hiroaki Ushida are co–corresponding authors. Ai Fujita Sajiki is the primary corresponding author and is responsible for sample analysis. Hiroaki Ushida is responsible for clinical information. This study was supported by a grant from the Japan Society for the Promotion of Science KAKENHI, grant number 20K09765 (Koji M Nishiguchi), 22K20958 (Ai Fujita Sajiki).

## Figure Legends

**eFigure.**
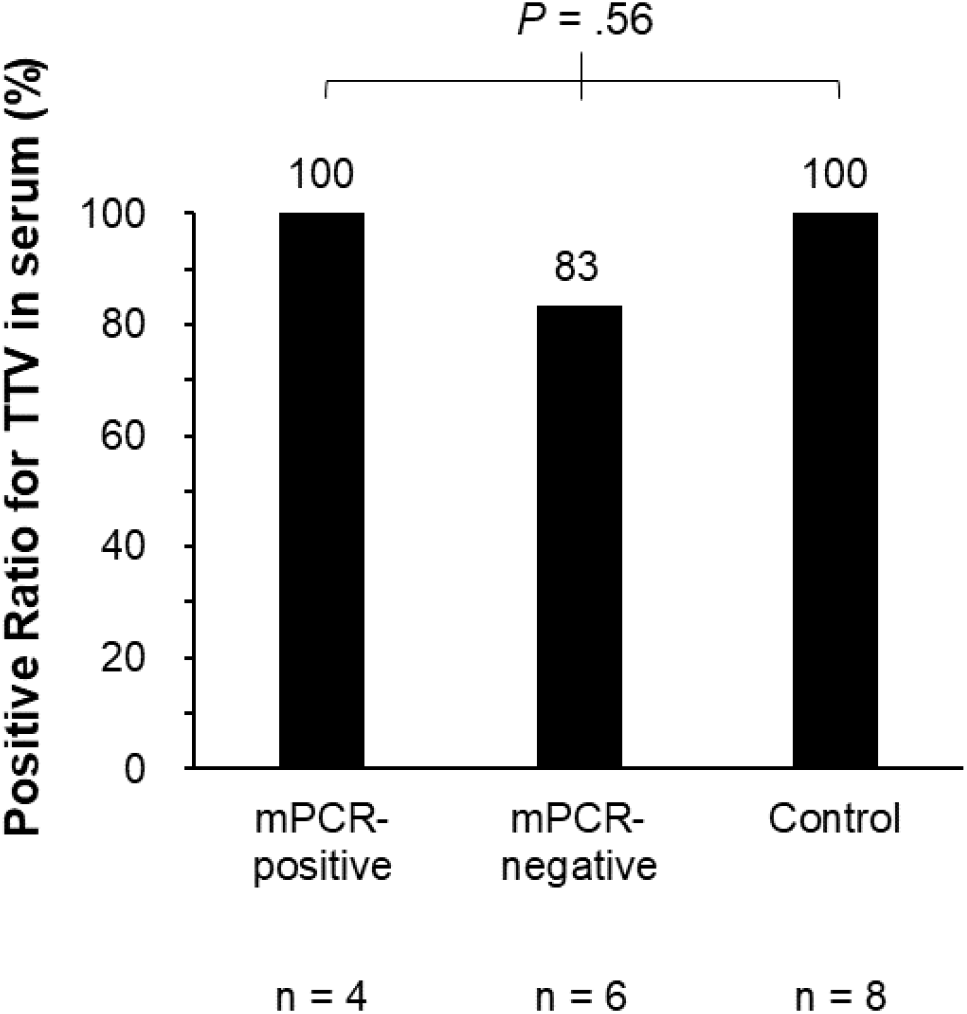
The disease-specific positive ratio for torque teno virus (TTV) in serum. A total of 18 samples of serum were collected at the same time as the AH samples. Each bar graph shows the positive ratio for serum TTV in each group. The Fisher–Freeman–Halton exact test was used to calculate *P* values, with *P* < .017 defined as significant after Bonferroni correction. Abbreviations: mPCR, multiplex polymerase chain reaction; TTV, torque teno virus.

**eTable 1.**
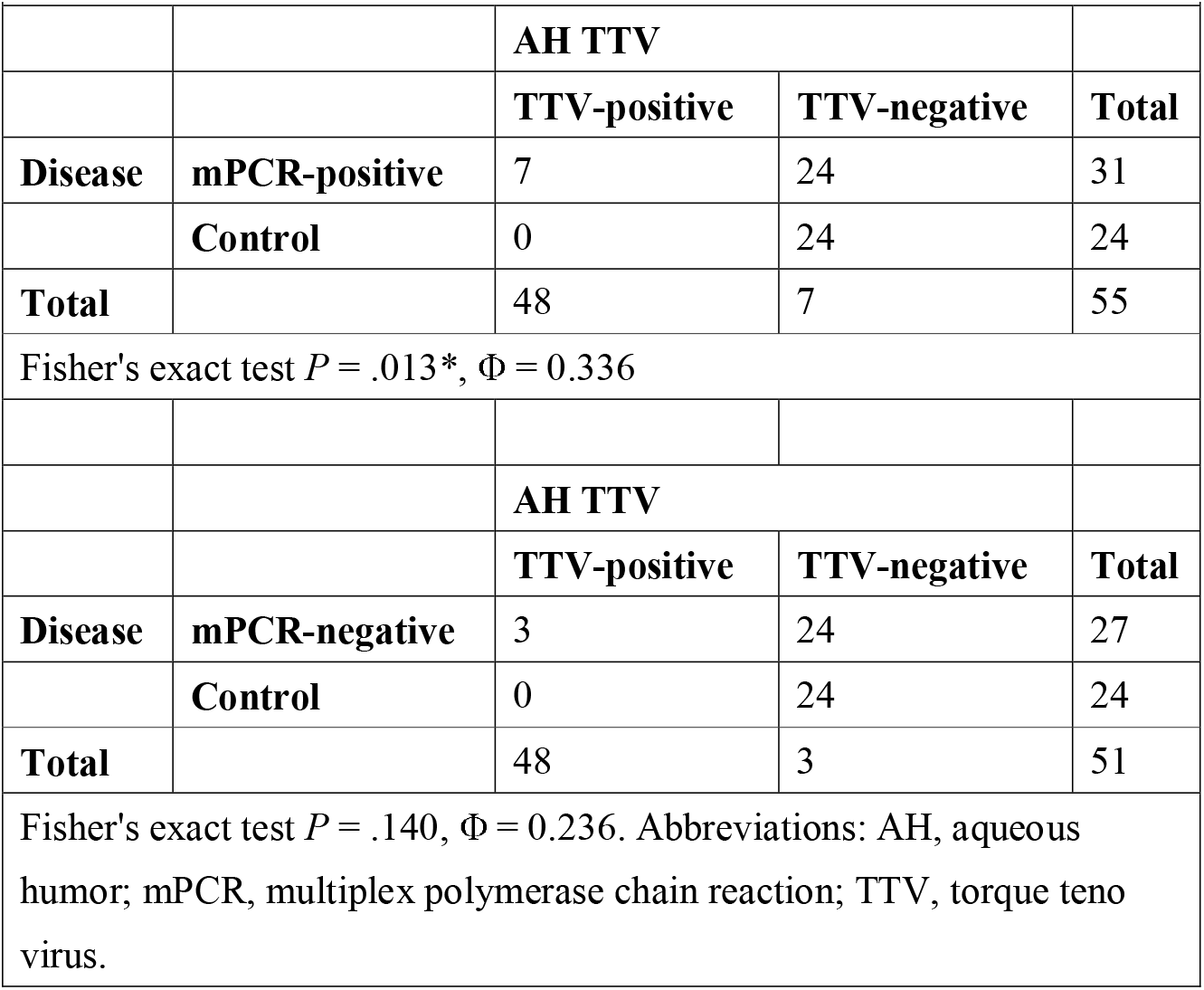
Cross Table Comparing the Detection of AH TTV in Patients With Different mPCR Results and Controls (Corresponds to Figure 1A)

**eTable 2.**
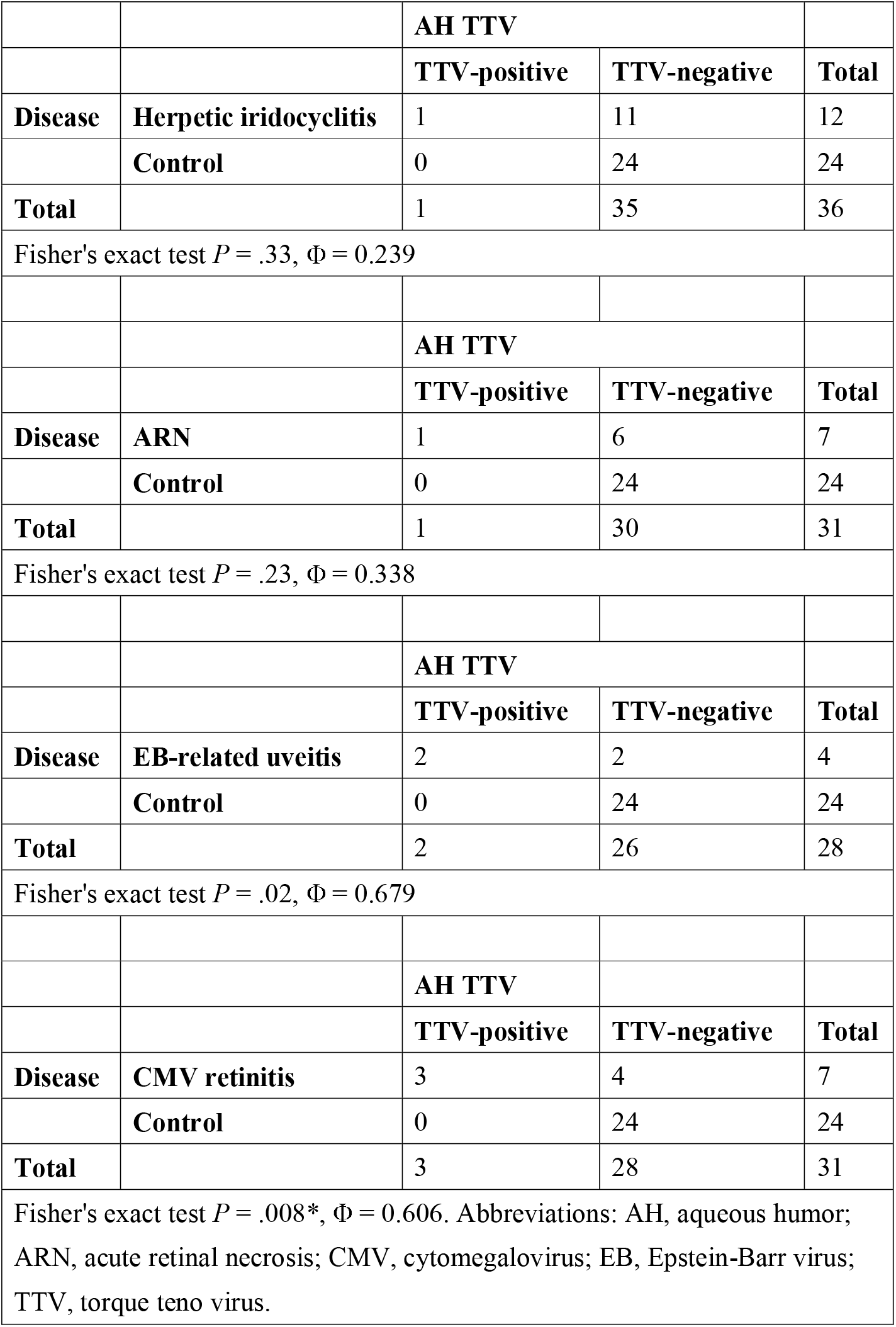
Cross Table Comparing the Detection of AH TTV in Different Types of Uveitis and Controls (Corresponds to Figure 1B)

**eTable 3.**
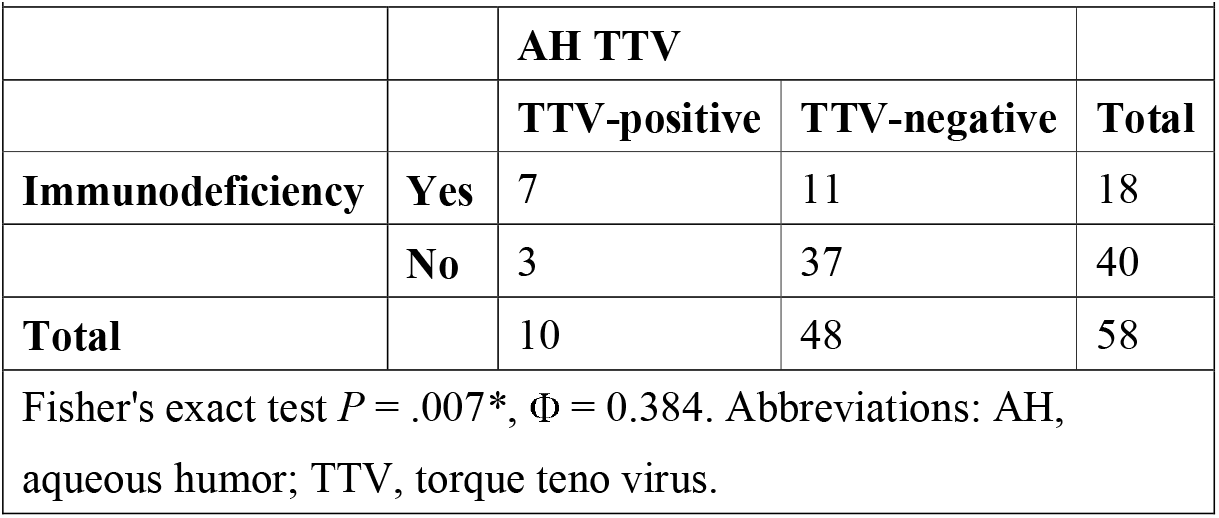
Cross Table Comparing the Detection of AH TTV and the Presence of Immunodeficiency (Corresponds to Figure 2)

**eTable 4.**
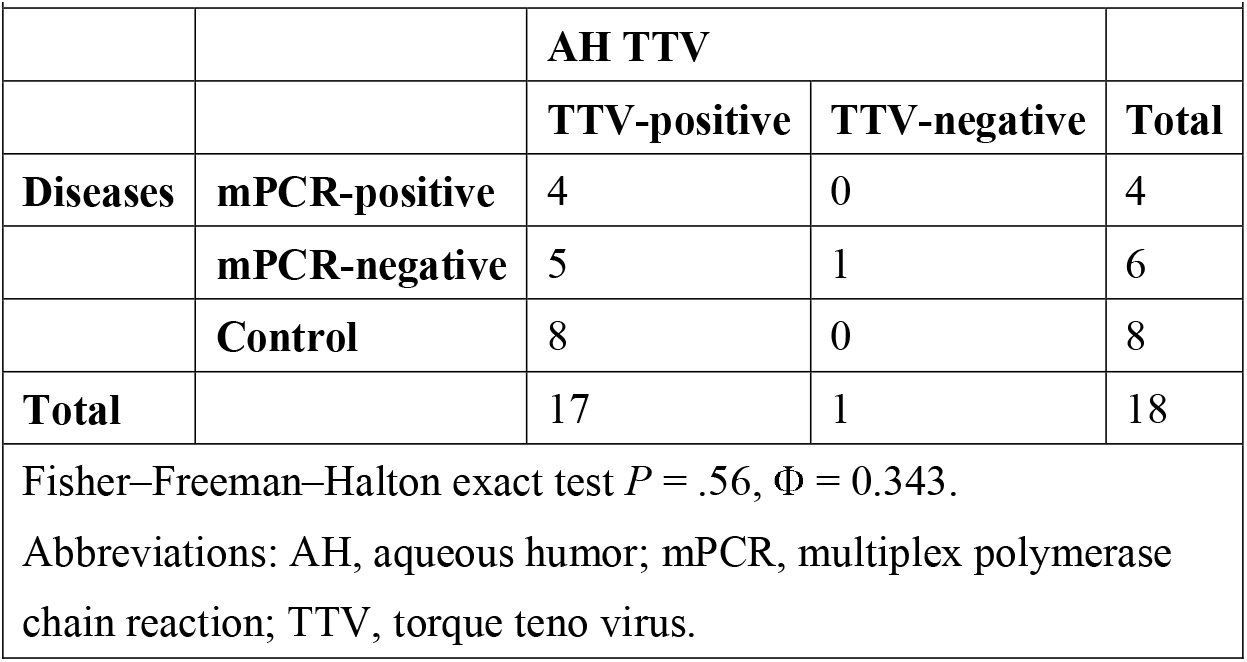
Cross Table Comparing the Detection of AH TTV and the Presence of Immunodeficiency (Corresponds to Figure 3)

